# Managing Data Uncertainty and Machine Learning for Adult ADHD Classification Using Accelerometry: OBF-Psychiatric Case Study

**DOI:** 10.1101/2025.08.26.25332257

**Authors:** Paola A. Castillo-Gutierrez

## Abstract

This study aims to enhance our understanding of ADHD individuals through accelerometer analysis while developing a framework for managing data uncertainty in digital biomarker research. Our primary emphasis is on identifying and mitigating biases within the OBF-Psychiatric dataset for ADHD and CONTROL groups, exploring how these biases influence machine learning model performance and generalizability. To balance patient inclusion with data quality, we applied innovative Pareto optimization and rigorous quality criteria, establishing optimal temporal windows, selecting 16:00 to 23:00 hours involving 53 patients from 77 available. Statistical analysis used robust Brunner-Munzel tests with False Discovery Rate correction, examining 34 comprehensive features spanning statistical, complexity, and frequency-based domains. Following thorough corrections, no significant differences in motor activity features emerged between the ADHD and CONTROL groups in quality-controlled data. Multidimensional scaling confirmed considerable overlap between groups. We assessed six traditional supervised machine learning algorithms through Leave-One-Patient-Out cross-validation: Logistic Regression, Random Forest, Support Vector Machine, Multilayer Perceptron, K-Nearest Neighbors, and XGBoost, plus baseline classifiers. Performance was evaluated across three data quality configurations to assess data processing consequences. Notably, performance systematically declined as data quality improved, with ROC AUC dropping from 75% (uncleaned) to 54% (quality-controlled). Our analysis suggests that the ADHD and CONTROL groups are indistinguishable using traditional feature-based motor activity patterns with these data collected in naturalistic conditions. However, the methodological contributions of this study provide foundations for future appropriately powered research and underscore key considerations for accelerometry utility in detecting adult ADHD and other psychiatric disorders in real-world scenarios.

## Introduction

Attention Deficit Hyperactivity Disorder (ADHD) is a neurodevelopmental disorder that affects approximately 5% of children globally [1] and is among the most common psychiatric disorders in childhood [2]. According to a meta-analysis conducted in 2020 [3], the adult ADHD landscape revealed two distinct prevalence patterns: the proportion of persistent cases (individuals who maintained their childhood diagnosis into adulthood) stood at 2.58%, while the rate of adult-onset cases (those who exhibited symptoms in adulthood without a childhood diagnosis) reached 6.76%, corresponding to 139.84 million and 366.33 million affected individuals worldwide, respectively. However, these figures may be imprecise due to evolving diagnostic criteria in adults, the historical underrepresentation of females in research, and the persistent stigma surrounding the disorder [4] [5]. Norway presents an ideal research context due to its universal and free healthcare system, where the private sector is marginal [6]. This unified system effectively diminishes variations in healthcare access and insurance status as potential confounding factors in ADHD diagnosis patterns. An extensive registry-based analysis by Solberg et al., drawing data from four national Norwegian databases (The Medical Birth Registry, Prescription Database, Patient Registry, and National Educational Database), investigated gender-specific patterns of psychiatric comorbidity in a cohort of 40,000 adults with ADHD [7]. The results demonstrated marked gender differences in comorbidity patterns: women exhibited significantly higher prevalence rates for anxiety, depression, bipolar disorder, and personality disorders, while men showed elevated rates of schizophrenia and substance use disorder [7].

Traditionally, licensed clinicians diagnose ADHD in adults primarily through structured clinical interviews, retrospective assessment of childhood symptoms, and application of diagnostic guidelines, such as the DSM-5, which focuses on manifestations of inattention, hyperactivity, and impulsivity [8] [5]. While this approach has effectively identified many cases, it remains limited by its reliance on subjective reporting and recall bias [4]. Furthermore, the substantial symptom overlap between ADHD and other common psychiatric disorders in adulthood can significantly impair clinicians’ diagnostic accuracy, leading to misdiagnosis and delayed treatment interventions [9] [4] [6].

Recent scientific findings have enhanced the identification standards for ADHD, although the evaluation instruments and assessment methodologies have stayed fundamentally the same [10] [5]. Significantly, the evolved data-driven and theoretical models designed to elucidate how neurodevelopmental disorders present across multiple dimensions have yet to be integrated into experimental research and everyday clinical practice [11] [12]. Most studies rely on magnetic resonance imaging (MRI) data to characterize the brain structure of individuals with ADHD, making it the primary quantitative method for comparison in research literature, thus far [10] [13]. The MRI procedures require expensive equipment, specialized facilities, and considerable time investments, making them impractical for routine diagnostic procedures. Additionally, MRI environments can be particularly challenging for individuals with ADHD symptoms, as these settings demand extended periods of stillness and can provoke anxiety [14].

In contrast, physiological signals such as motion data from accelerometers and inter-beat intervals, gathered with electrocardiography or plethysmography, offer more accessible and costeffective alternatives. Researchers collect these biomedical signals in naturalistic settings, with minimal patient cooperation required, while enabling continuous monitoring capabilities [15]. Despite the advantages, a considerable gap remains in understanding the potential and constraints of accelerometry as an alternative to biomedical signals, particularly in distinguishing ADHD within the adult population [16, 13]. This knowledge gap is especially striking given the increasing demand for objective, scalable diagnostic and prognostic tools that healthcare professionals readily integrate into diverse clinical environments. Successful machine learning studies require a clear clinical rationale, robust validation strategies, a detailed methodology that includes feature engineering and preprocessing, a considerable amount of data, and transparent reporting of limitations and generalizability constraints. This paper aims to enhance our understanding of accelerometry signals by characterizing the limitations researchers should thoroughly evaluate when using the ADHD and CONTROL data from the OBF-Psychiatric Dataset [17] to classify patients using machine learning.

The principal research question that guides this work is:

**To what extent can traditional supervised machine learning models, using accelerometer-based feature extraction on an aggregated daily basis, differentiate ADHD patients from a control group of adults for the OBF-Psychiatric selected dataset?**

For this problem and data, the secondary research question is:

*How does the three-way trade-off between maximizing patient inclusion, maximizing recording duration, and minimizing confounding factors affect the performance of accelerometry-based ADHD classification models?*

This work presents the following contributions:

- Comprehensive analysis of the ADHD and CONTROL data incorporated within the OBF-Psychiatric dataset [17], including detailed characterization of the inherent properties of both accelerometer measurements and clinical variables.
- A rigorous quality assessment of the acquired accelerometry signals, alongside the development of customized exclusion criteria specifically designed to address data acquisition anomalies in the samples. To ensure analytical consistency across patients and diagnostic classifications, we identified time windows using a Pareto front approach, which determined the set of non-dominated solutions that balance patient inclusion and window duration. This application could be transferable to other accelerometry or time series cases where quality constraints restrict the generalizable window across several patients. The resulting curated dataset, containing records from both the CONTROL and ADHD groups, has been made publicly accessible through Zenodo to facilitate further research in this domain [18].
- Comparison of basic and complex features extracted from accelerometer data across various time-frame selection approaches using inferential statistical modeling.
- Development and evaluation of binary machine learning classifiers for ADHD detection based on accelerometry data, with performance metrics systematically assessed at the patient level across three carefully selected temporal configurations:
  a. Analysis incorporating all available complete-day recordings (00:00-23:59) from all patients (n=77) without additional quality filtering.
  b. Analysis using the Pareto-optimal window (16:00-23:00) that maximizes patient inclusion (n=53) while maintaining data quality criteria and minimizing Euclidean distance to a “Utopia point” established at (77 patients, 24 hours).
  c. Full 24-hour analysis (00:00-23:59) using the same 53 patients identified through the Pareto-optimal window criteria.

The structure of this paper is organized as follows: Section II provides a literature review of relevant studies related to this research. Section III outlines the methodology utilized. Section IV presents the results derived from the machine learning and statistical inference experiments, along with a discussion of the findings. Section V addresses several limitations and special considerations to be aware of when handling these data. Finally, Section VI summarizes the work and identifies potential future research directions and opportunities.

### Literature Review

#### Neurobiology and Clinical Aspects of ADHD

The presentation of ADHD varies considerably across individuals, reflecting diverse developmental trajectories and manifestation patterns; rather than being a binary manifestation of symptoms, it was suggested as a spectrum [19]. As a psychiatric condition, ADHD continues to be the subject of extensive research aimed at better defining its boundaries with other disorders and understanding its heterogeneous nature. However, despite these efforts, the fundamental causes and developmental pathways that give rise to ADHD continue to elude complete scientific explanation [20].

Various psychological frameworks have been proposed to explain the origins of symptomatic hyperactivity and motor activity in ADHD, building upon foundational neuropsychological theories that emphasize executive dysfunction and inhibitory control deficits [21, 22]. These include: (1) the State Regulation Model, which examines arousal and activation mechanisms; (2) Multiple Pathway Theories, which consider diverse developmental routes to the disorder; and (3) the Dynamic Developmental Theory of ADHD, which focuses on altered reinforcement processes [14]. The Dynamic Developmental Theory of ADHD attributes hyperactivity and motor symptoms to a hypo-functioning mesolimbic dopamine branch, resulting in altered reinforcement of new behaviors and inadequate extinction of previously reinforced behaviors [23]. This theory predicts that hyperactivity develops gradually through a combination of disrupted positive reinforcement and deficient extinction processes, ultimately manifesting as excessive motor activity from an accumulation of behavioral responses that cannot be adequately regulated due to narrowed time windows for associating antecedent stimuli and behavior with consequences [14, 23, 24]. Both DSM-IV and DSM-5 explicitly note that adults may “feel restless” instead of running or climbing, and both require symptoms in two or more settings, underscoring that motor hyperactivity can be situational [25] [8]. Hyperactivity symptoms generally diminish with age [26]. Table I summarizes the main differences between DSM-IV and DSM-5, describing the presentation of hyperactivity and motor symptoms, along with other factors such as age of onset. The DSM-IV [25] and ICD-10 [27] are two systems that share similar lists of symptoms, with one key difference being that the DSM-IV explicitly recognizes distinct subtypes, including predominantly inattentive presentation, whereas ICD-10 employs a more unified approach to hyperkinetic disorders. The ICD-10 hyperkinetic disorder (HDK) does not formally recognize ADHD, but resembles the combined presentation ADHD type of ADHD because there are symptoms of inattention and hyperactivity/impulsivity.

**Table 1.** Comparison of ADHD Diagnostic Criteria Between DSM-IV (1994) [25] and DSM-5 (2013) [8].

| Criterion | DSM-IV (1994) | DSM-5 (2013) |
| --- | --- | --- |
| **Motor Symptoms:** |  |  |
| Fidgeting / Squirming | "Often fidgets with hands or feet or squirms in seat." | "Often fidgets with or taps hands or feet, or squirms in seat." |
| Leaving Seat | "Often leaves seat in situations when remaining seated is expected." | "Often leaves seat in situations when remaining seated is expected." |
| Running / Climbing | "Often runs about or climbs excessively in situations in which it is inappropriate (in adolescents or adults, may be limited to subjective feelings of restlessness)." | "Often runs about or climbs in situations where it is not appropriate (adolescents or adults may be limited to feeling restless)." |
| "Motor-Driven" Behavior | "Is often 'on the go' or often acts as if 'driven by a motor'." | "Is often 'on the go' acting as if 'driven by a motor'." |
| **Key Diagnostic Changes:** |  |  |
| Age of Onset | "Symptoms that caused impairment were present before age 7 years." | "Several inattentive or hyperactive-impulsive symptoms were present prior to age 12." |
| Number of Symptoms Required (Adults 17+) | "Six or more symptoms required for all ages". No special distinction for older adolescents or adults. | "Five or more symptoms required for adolescents 17+ and adults. Required 'presence of', rather than 'impairment by' symptoms." |
| Classification | "Classified under 'Disorders Usually First Diagnosed in Infancy, Childhood, or Adolescence'." | "Reclassified under 'Neurodevelopmental Disorders'." |

The symptom profile of ADHD extends beyond traditional conceptualizations of attention deficits and hyperactivity, encompassing complex patterns of cognitive and behavioral dysregulation. Individuals with ADHD often experience intense fluctuations in attention capacity, demonstrating the ability to hyperfocus on engaging tasks while struggling with sustained attention in other contexts. This pattern suggests an underlying dysregulation of attention mechanisms rather than a simple deficit [28]. Attention deficits in ADHD may result from insufficient dopamine activity in the prefrontal cortex, a hypothesis supported by the therapeutic efficacy of stimulant medications that enhance dopaminergic function [29]. Executive function challenges manifest in difficulties with time management, organization, and task completion. Clinical evidence suggests adults experience a broader range of impairments than children, particularly in executive functioning and emotional regulation [30, 4]. Despite the challenges they face, individuals with ADHD often possess remarkable strengths that can contribute to their success in adulthood. They frequently demonstrate exceptional creativity, boundless curiosity, and natural problem-solving abilities. Their characteristic energy, nonconformist thinking, adventurous spirit, spontaneity, and keen sense of humor represent valuable attributes that, when properly channeled, can become significant advantages in appropriate contexts and career paths [19].

#### Accelerometry Signals

Physical activity is a complex human behavior that requires multiple dimensions to be considered in order to reveal a comprehensive behavioral profile. The psychological factors of cognitive and emotional function reflect the desire to maintain productivity, independence, and an active interaction (movement) within their environment [31]. Accelerometry has emerged as the most commonly used and evaluated objective method in clinical and epidemiological research to capture movement, offering a solution to the limitations of subjective measurement approaches [32].

Modern accelerometers consist of small sensors that register acceleration along specific axes and can be worn at various body locations, with the hip, wrist, and thigh being the most common placements. Accelerometers provide a convenient and unobtrusive method for continuous data collection. Multiple accelerometers are used in research and consumer devices, including piezoelectric, piezoresistive, and capacitive accelerometers, each utilizing different mechanisms to detect acceleration forces [31]. Accelerometer signals can be obtained either as raw acceleration data measured in gravitational units (g) or as processed activity counts after applying algorithms to the raw signals.

The medical-grade Actiwatch AW4 device, specifically, employs a piezoelectric element with a seismic mass that detects movement above a threshold of 0.05g in multiple directions (anteriorposterior, mediolateral, and vertical planes), with a primary focus on the vertical axis (omniaxial type), converting mechanical acceleration into digital signals. Frequency filters are set to a range of 3 to 11 Hz. This device samples acceleration at a predefined 32 Hz frequency, selects the highest sample value in one second and ultimately transforms it into activity counts using an algorithm where 128 counts per second corresponds to approximately an acceleration of 5g (which is also considered as a maximum for human acceleration typical range in the device manual) [33]. These counts are then summed and stored in predefined epoch lengths (commonly 15, 30, or 60 seconds) to quantify movement intensity over time. The device manual indicates that it is not suitable to use underwater, as this could damage the accelerometer [33].

According to the Nyquist theorem, to accurately capture a signal, the sampling rate must be at least twice the frequency of the signal being measured. Human movement acceleration signals predominantly occur below 10 Hz, suggesting that even relatively modest sampling rates are adequate for capturing the full spectrum of human movement patterns [34]. This presents an important trade-off in accelerometer design, as higher sampling rates and resolutions provide more detailed data but require significantly more memory and power; however, this is increasingly becoming less of a concern with ongoing technological development. For typical human movement monitoring, an accelerometer with a 2G measurement capability is generally sufficient [34]. However, the accelerometry device domain presents substantial challenges for researchers attempting to compare results across studies or establish universal clinical findings. The commercial market features accelerometers with widely varying technical specifications, including hardware-level filtering mechanisms, sampling frequencies, output metrics (such as activity counts or raw acceleration), and proprietary analytical algorithms that manufacturers protect as intellectual property. These technical inconsistencies, along with differences in clinical protocols for device placement, epoch durations, and non-wear-time imputation algorithms, create significant confounding factors when comparing digital biomarkers derived from different accelerometry systems [35]. When utilizing the same accelerometer model across a study, researchers should still conduct precision and accuracy testing on each sensor unit before data collection to minimize device-specific variance and systematically document the calibration procedure or establish a criterion value [36, 35, 34].

It is recommended to adopt a seven-day analysis period in clinical protocols for accelerometry-based physical activity assessment, as this duration strikes a balance between data reliability and participant compliance while capturing sufficient behavioral variability across weekdays and weekends [32]. Designing the post-processing techniques for these signals requires knowledge of biomechanics, psychology, physiology, engineering, computer science, and statistics. These multidisciplinary considerations play a pivotal role in ensuring accurate measurement with accelerometers and valid interpretations.

#### Related Works

The HYPERAKTIV dataset [15], which is now part of the OBF-Psychiatric dataset, has established itself as a foundational resource for ADHD research utilizing wearable sensors and motor activity data from adults, representing one of the few publicly available repositories of this kind [15]. The analytical approaches applied to these actigraphy measurements have consistently employed binary classification frameworks to distinguish ADHD from defined groups, with multiple studies augmenting the control group by combining clinical subjects with control individuals from complementary datasets, such as PSYKOSE and DEPRESJON [15, 37, 38, 17, 39]. Feature extraction has employed diverse methodologies across studies, ranging from traditional statistical features to automated tools like the Tsfresh Python library [40] emerging as the predominant tool for generating statistical features from time series data, followed by various dimensionality reduction and feature selection techniques including PCA [41, 37], UMAP, and t-SNE [38]. Validation methodologies have varied across studies, with researchers employing K-fold cross-validation [41] and percentage splits of training/testing data [15, 37]. In some cases, leave-one-out cross-validation [42] is used. Recent innovations by Thelagathoti and Ali [43, 44] employing network-based approaches represent promising steps toward more interpretable models. The reported classification accuracies using these data to date range from 72% [15] to 99.2% [38].

In Fasmer’s earlier 2015 study [45], researchers reported experiencing logistical issues and patient compliance problems during the study. They utilized a more restricted sample of only 32 ADHD patients and 20 CONTROL subjects after data quality inspection for six days. Their analysis excluded recordings containing daytime (07:00-23:00) inactivity periods exceeding one hour from the six-day analysis, but found no significant differences between the ADHD, CONTROL, and CLINICAL groups using activity counts for one half hour as the unit of measurement. In Fasmer’s 2020 diurnal graph theory study of these ADHD patients [46], researchers employed a methodology that analyzed only the first continuous 24-hour period (00:00-23:59) without data gaps for each participant. Within this timeframe, they focused on 6-hour morning periods (8:00 AM to 2:00 PM) and evening periods (6:00 PM to midnight), explicitly excluding the midnight to 8:00 AM interval from their analysis. The number of subjects considered in the 2020 study varied across measurements, with 30 control subjects and 40-42 for ADHD patients. Both the 2015 and 2020 studies mention that participants were instructed to wear actigraphs at all times and only remove them during showering or bathing.

While previous works demonstrate the potential of machine learning approaches for ADHD classification using accelerometry data, additional considerations regarding assumptions, data preprocessing steps, signal noise handling, and evaluation could further strengthen advancements in the field. To our knowledge, this study represents the first comprehensive approach to addressing these considerations, utilizing the ADHD and CONTROL data from the OBF-Psychiatric dataset and machine learning techniques.

### Methodology

We implemented multi-objective Pareto optimization to identify optimal temporal windows that balance patient inclusion with data quality constraints. The analysis included rigorous data preprocessing with custom quality criteria, extraction of 34 statistical and complexity features from accelerometer signals, and evaluation of six traditional machine learning algorithms using leave-one-patient-out cross-validation. Statistical inference was conducted using non-parametric tests with False Discovery Rate correction, while model performance was assessed across three distinct data quality configurations to evaluate the impact of preprocessing decisions on classification outcomes.

### Data Collection

The ADHD data encompasses data collected from 103 patients referred to a private psychiatric outpatient clinic in Norway who required diagnostic evaluation for various psychiatric conditions, specifically attention deficit/hyperactivity disorder (ADHD), mood disorders, or anxiety disorders. Data collection commenced in February 20, 2009. Diagnostic assessments for all participants were conducted by experienced and certified psychiatrists, utilizing the Mini-International Neuropsychiatric Interview (MINI Plus, version 5.0) [47], which was enhanced when possible with data from collateral sources, particularly relatives, concerning symptoms of ADHD in childhood [48]. A consensus final diagnosis was established using the DSM-IV and ICD-10 criteria, following thorough discussions of each case among various psychiatrists [48] [17]. Among these new referral patients, 51 were diagnosed with ADHD; however, motor activity data were recorded for only 45 of the ADHD patients (24 males and 21 females). This data collection occurred under naturalistic conditions, allowing participants to engage in their typical daily activities while wearing an accelerometer. The study incorporated inclusive criteria, accommodating comorbidities frequently observed in conjunction with substance abuse. Patients diagnosed with ADHD presented additional psychiatric conditions such as anxiety, bipolar disorder, or unipolar depression, resulting in substantial overlap with other psychiatric disorders. Additionally, the dataset features a category labeled “OTHER,” which denotes the presence of additional unspecified psychiatric disorders. The ages of participants ranged from 18 to 65 years. Concerning medication status, the majority of participants were unmedicated at the time of data collection, with only one individual within the ADHD cohort receiving a prescription for stimulants, but some receiving antidepressants, anxiolytics/benzodiazepines, and sleep medications (hypnotics) [45]. Participants maintained their medication regimen unchanged throughout the actigraphy monitoring period, continuing to take the same drugs they were using at the time of referral [48]. All collected data were anonymized to protect patient confidentiality, with each participant identified only by a unique numeric ID.

The CONTROL group data comprised 32 individuals (20 females and 12 males) with an age range between 21 and 66 years, recruited as part of the original DEPRESJON and PSYKOSE datasets, with data collection for this group beginning on October 2, 2002. This cohort included 23 employees from Bergen University and a psychiatric nursing home, five medical students, and four general practitioner patients who had no psychiatric history. No individuals in the control group had any previous diagnosis of mood disorders or psychotic conditions. Similar to the HYPERAKTIV dataset, accelerometry for the control group was performed under naturalistic conditions.

For this analysis, only patients diagnosed with ADHD and individuals from the CONTROL group from the OBF-Psychiatric dataset were selected for inclusion, despite the presence of patients in the CLINICAL group with accelerometry recorded in naturalistic settings. This decision was made to reduce heterogeneity and clarify the classification structure, as the CLINICAL group encompassed individuals with a variety of diagnoses and comorbidities. While the DEPRESSION and SCHIZOPHRENIA groups from the PSYKOSE and DEPRESJON datasets were available, they were not included due to their differing protocol descriptions. The SCHIZOPHRENIA group consisted entirely of hospitalized patients who were not working, predominantly male, which raises concerns about the underrepresentation of females and potential fallacious results when using this group. Meanwhile, the DEPRESSION group included both hospitalized and outpatient individuals. In parallel, this approach allows for a preliminary assessment of whether meaningful differences exist between individuals with ADHD and those without psychiatric conditions in naturalistic conditions. Accelerometer data were recorded for both groups using the Actiwatch AW4 on the right wrist. Exploratory visuals of medication distribution and demographics from these groups, such as gender, age, and psychiatric comorbidities, are included in the repository [18].

The original protocols for data collection from the studies received approval from the Norwegian Regional Medical Research Ethics Committee West, and their specific numbers are included in the OBF-Psychiatric data descriptor paper [17].

### Data Cleaning and Preprocessing

While the literature suggests analyzing seven consecutive days, this was not feasible due to the limitations of the available data. The number of patients in the ADHD group who met this criterion dramatically declined from 45 to just 6, significantly restricting our analytical capabilities.

The activity data was structured into matrices for analytical clarity, with each matrix representing complete 24-hour periods (1,440 minutes from 00:00 to 23:59) as individual vectors. For the CONTROL group, there were even more days recorded than those reported in the clinical study, extending to weeks. We decided to consider only the maximum number of days documented in the original.csv file for this group, as established in the “days” column, by considering the full days available and skipping the first incomplete day. For the ADHD group, 45 patients were found, totaling 253 complete days, while the CONTROL group consisted of 32 patients, totaling 402 complete days.

We graphed the motor activity for each patient across daily segments and visually identified several signal acquisition issues; examples of these are shown in Figure 1. Since it remains unclear whether participants consistently wore the watch during both nighttime and daytime or periodically removed it, we searched for an interval between 7:00 and 23:00 hours that could be generalizable across patients and classes, considering the following quality criteria:

**Figure 1.**
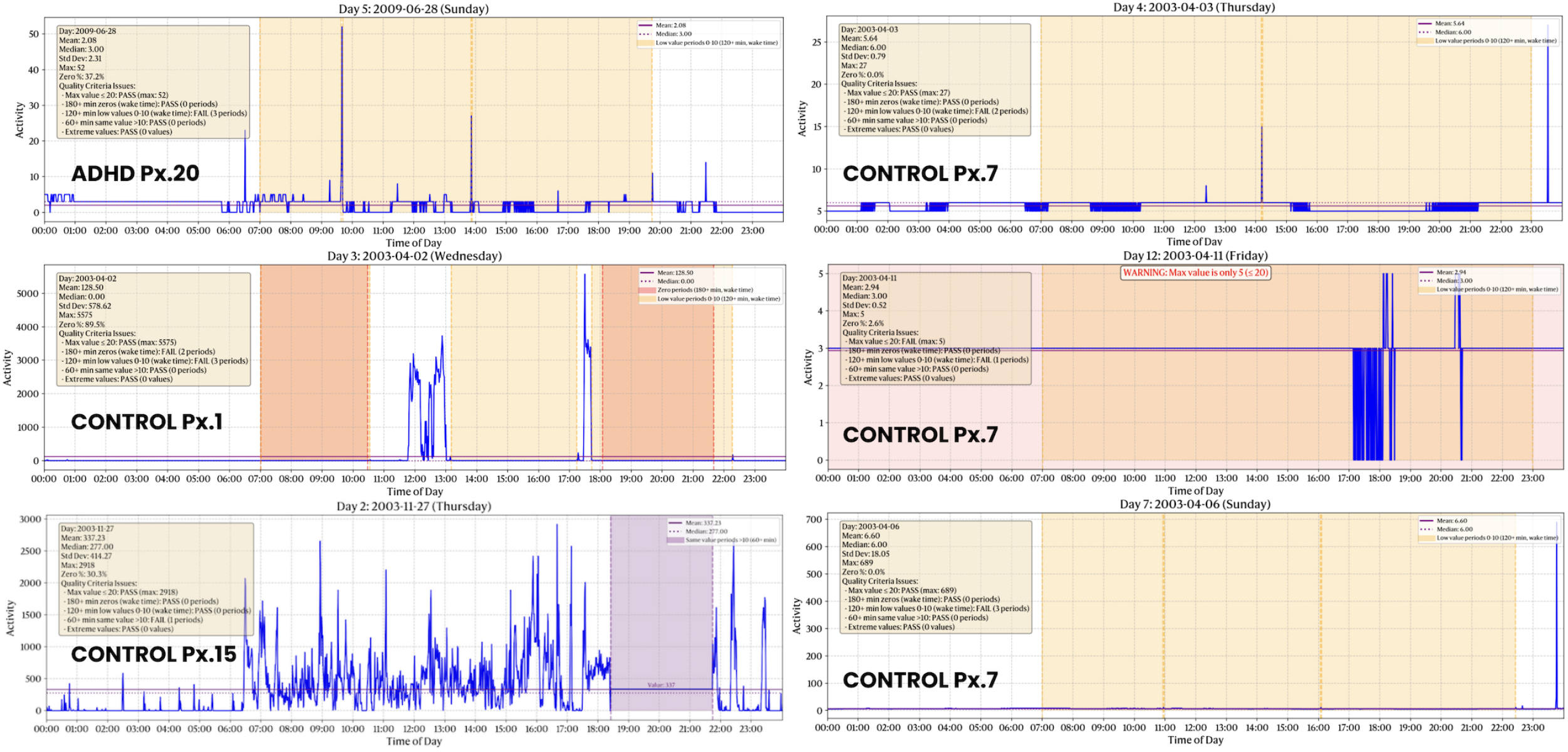
Examples of signals with acquisition anomalies.

1. No day segment should have 180+ consecutive minutes with zero values during presumed wake time (07:00-23:00).
2. No day segment should have 120+ consecutive minutes with values between 0-10 during presumed wake time (07:00-23:00).
3. No day segment should have 60+ consecutive minutes with the same value (if that value is > 10).
4. No day segment should contain extreme values between 7680 and 8000.
5. No day segment should contain more than 70% of zeros during the day segment.
6. No day segment should contain only a maximum value of 20 counts or less.
7. Each window must have at least 5 hours of data, and each patient should have six consecutive days available.

We applied the principles of Pareto optimality to identify a set of non-dominated solutions that optimize both patient inclusion and window duration. An exhaustive search of the Pareto front examined all possible time windows from 7:00 to 23:00, with a minimum window size of 5 hours. For each potential time window, we evaluated the number of patients who met the quality criteria along with the corresponding window duration. A solution was considered Pareto-optimal if no other window could simultaneously include more patients and span a longer duration. Let *p* (*w*) represent the number of patients meeting quality criteria for window *w*, and *D*(*w*) represent the duration of window *w* in hours. A window *w*_1_ dominates another window *w*_2_ if and only if:

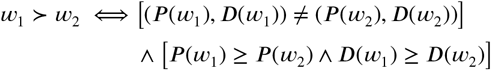

Genetic Algorithms and other heuristic methods are commonly used to identify Pareto-optimal solutions within large search spaces. However, the specific dimensionality of our problem permitted a comprehensive exploration of the entire solution space, ensuring the accurate identification of the actual Pareto front rather than merely an approximation. We defined a “Utopia point”, specifically at (77 patients, 24 hours), as the ideal maximum in the absence of confounding factors. This point served as a reference for calculating Euclidean distance to identify the solution from the Pareto Front that minimizes this distance. Our exhaustive search identified six non-dominated solutions forming the Pareto front (see Figure 2). From this set, we selected only one key window for detailed analysis:

**Figure 2.**
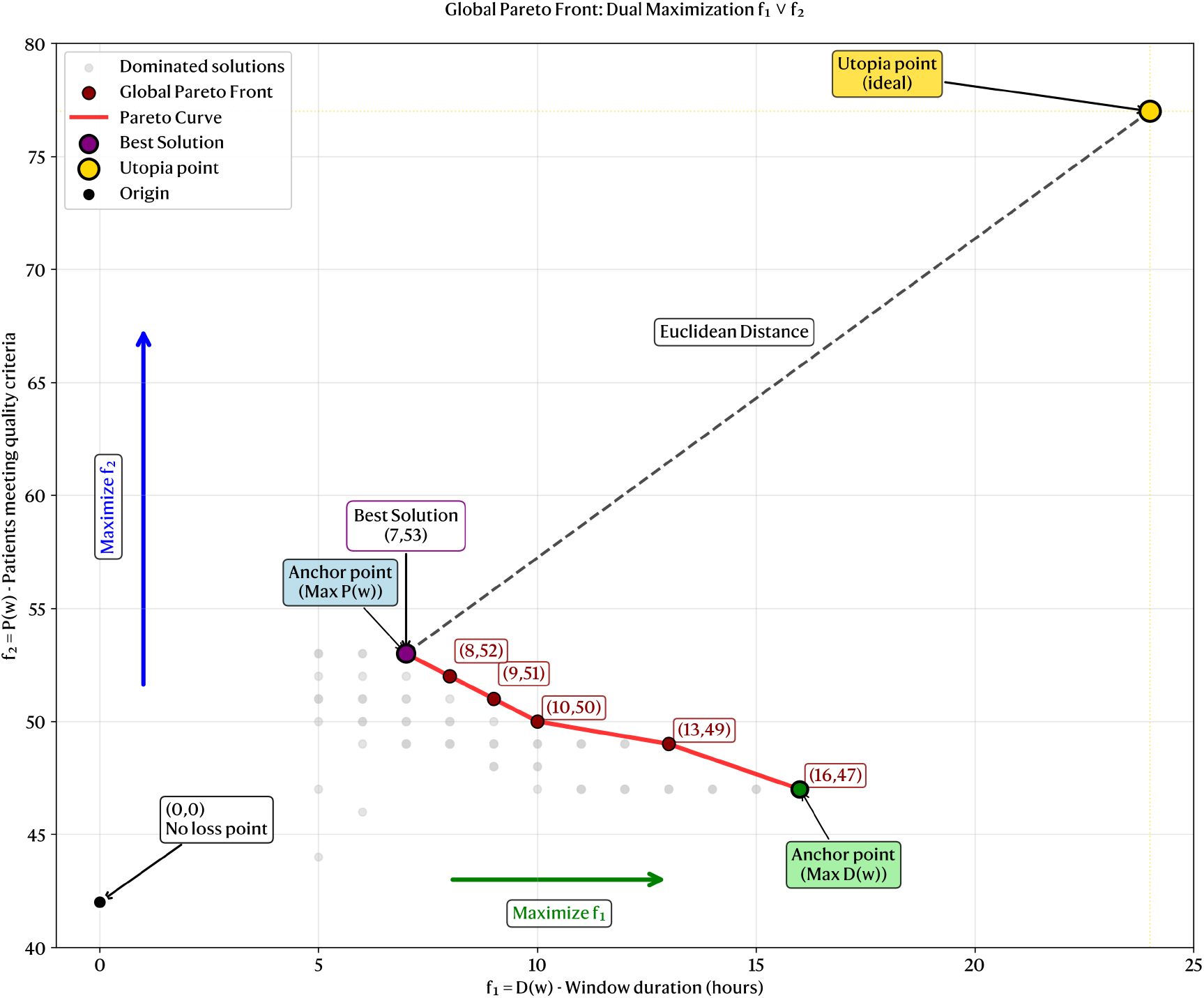
Global Pareto Front for Time Window Optimization in Actigraphy Data Analysis.

### 1. Optimal Window (maximizing patient inclusion and minimizing Euclidean distance)

16:00-23:00 (7 hours), which included 53 patients (28 ADHD and 25 CONTROL)

The ADHD group had no more than 7 days available, whereas the CONTROL group did; however, we only considered the first 6-day quality sequence for the CONTROL group. To ensure robust findings and evaluate the impact of different temporal selection criteria, we extended our analysis beyond the Pareto-optimal windows to include two additional temporal configurations. First, we conducted an analysis that incorporated all available complete-day recordings (00:00-23:59) without additional quality filtering, thereby maximizing cohort size at the expense of potentially including lower-quality segments. Second, we performed a complete 24-hour analysis (00:00-23:59) using the same 53 patients and six days identified through the Pareto-optimal window criteria (16:00-23:00). This multi-configuration approach facilitated the evaluation of classifier performance sensitivity to temporal window selection and quality filtering parameters. The processed actigraph recording files, adapted to each Pareto window configuration, are available in the Zenodo repository [18], including both “TRIMMED” (Pareto-window-specific) and “COMPLETE” (full 24-hour recordings for the same patients) versions.

### Feature Extraction

We extracted nine core basic features per day segment for each patient: mean, standard deviation, median, percentage of zeros (minutes with value 0 for activity), quartile 25, quartile 75, percentile 95, skewness, and kurtosis. Additionally, an extended set of 25 complementary features was calculated to capture comprehensive activity patterns: Median absolute deviation (MAD), 10% trimmed mean, amplitude, percentage of minutes in ranges 1–100, 101–500, 501–1000, 1001–2000 counts, percentage of low activity values ≤ 40, M60 (mean of 60 highest activity values), intensity gradient, activity above mean percentage, activity variability ratio, Gini coefficient, activity fragmentation index, heavy tail ratio (P99/P75), extreme proportion (>P90 threshold), Detrended Fluctuation Analysis (DFA) scaling exponent, high frequency power (1/30 to 1/2 cycles per minute), low frequency power (1/210 to 1/60 cycles per minute), HF/LF ratio, sample entropy with embedding dimension m = 2 and tolerance r = 0.2× standard deviation, normalized Lempel-Ziv complexity using median-based binarization, permutation Lempel-Ziv complexity with embedding dimension m = 3 and autocorrelation at 30 and 90-minute lags. The files with the daily and patient-level extracted features are available in the repository [18].

The normality of each feature distribution was assessed using the Shapiro-Wilk test, while homogeneity of variance was evaluated through Levene’s test to determine the appropriate statistical inference methodology. For this part, we aggregated the features by calculating the mean across all six days for each patient, as independence cannot be assumed between daily observations from the same individual; however, it can be reasonably established between different patients. Based on these preliminary analyses, the Brunner-Munzel Test was selected as the most appropriate non-parametric method for between-group comparisons. For comparison with results obtained using the Tsfresh library, the Mann-Whitney U Test was additionally included in the analysis. Multiple comparison correction was performed using the Benjamini-Hochberg false discovery rate (FDR) procedure with *α* = 0.05 to control the expected proportion of false positives among rejected hypotheses. We considered 34 comparisons because of the number of features. Statistical significance was established at p < 0.05. We calculated the effect sizes (Cohen’s d) to quantify the magnitude of differences. The 95% confidence intervals for the mean values of each group and each feature were estimated using bootstrap resampling with 1000 iterations. Feature-to-feature relationships among motor activity metrics were analyzed using Spearman’s rank correlation, a non-parametric method suitable for these data. Demographic effects on features were assessed through Kruskal-Wallis tests with FDR corrections across gender (4 subgroups: ADHD-female, ADHD-male, CONTROL-female, CONTROL-male) and age subgroups (6 subgroups across three age ranges: 17-34, 35-51, and 52-69 years per diagnostic group).

Multidimensional Scaling in three dimensions was used to see the classifying potential of the set of features at the daily level per class. We chose this dimension because it preserves the most information that could be lost through dimensionality reduction, while maintaining the visualization capabilities.

### Machine Learning Model Training and Validation

The traditional machine learning models chosen for evaluation were Logistic Regression (LR), Random Forest (RF), Support Vector Machine (SVM), Multilayer Perceptron (MLP), K-Nearest Neighbors (KNN), and XGBoost. Fixed hyperparameters and Python notebook code are available in the repository for reproducibility. Baseline classifiers with Most Frequent, Stratified, and Uniform strategies were implemented for benchmark comparison against traditional machine learning models. These baseline classifiers were trained using the overall class distribution from the complete patient dataset rather than following the leave-one-patient-out cross-validation scheme. This approach ensures stable baseline references that reflect the inherent class proportions in the data, avoiding the distribution distortions that could arise from small sample scenarios. The baseline classifiers operate at the patient level and provide predictions based solely on learned class frequencies, without requiring majority voting aggregation.

A leave-one-patient-out cross-validation approach was employed, wherein each patient’s data was systematically excluded in each fold to ensure robust model generalization. Within each fold, features were independently normalized using Min-Max scaling, applied on a per-feature basis to constrain values between 0 and 1 while preserving the relative distribution within each feature and transforming the test set at each iteration independently. Class imbalance was addressed through the application of the Synthetic Minority Over-sampling Technique (SMOTE) [49], applied exclusively to the training partition of each fold and with a random seed (42). Predictions were subsequently generated at the daily level (either complete period or segment), providing temporal granularity. The final label assigned for each patient is based on the majority voting of the daily predictions.

Model performance was evaluated using a comprehensive set of metrics: precision, recall, F1 score, Receiver Operating Characteristic Area Under the Curve (ROC AUC), accuracy, and Matthews Correlation Coefficient (MCC). These metrics were selected to provide a balanced assessment of classification performance across different dimensions of model evaluation.

### Intra-patient and inter-patient analyses

The intra-patient analyses examine behavioral dynamics within individual participants across time, encompassing two key dimensions: temporal correlations that quantify the relationship between each feature and time progression within each patient’s longitudinal data using Spearman correlation coefficients between time points and feature values; and variability analysis that captures the coefficient of variation

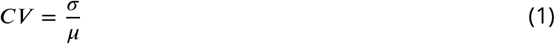

to identify features with high intra-individual fluctuation. Complementing these within-subject analyses, the inter-patient methods investigate patterns across individuals within each diagnostic group, focusing on consistency using intraclass correlation coefficients (ICC) calculated as

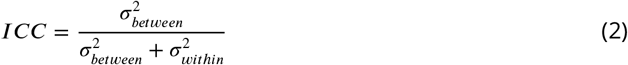

to quantify how homogeneous patient groups are in their feature expressions. These analyses were made only for the Pareto optimal window.

## Results and Discussion

Our implementation of multi-objective Pareto optimization for temporal window selection, shown in Figure 2, revealed methodological trade-offs. The exhaustive evaluation of 78 window combinations (51,090 individual assessments) identified six non-dominated solutions, with the optimal window (16:00-23:00, 7 hours, 53 patients, 68.8% inclusion) representing the best compromise between temporal coverage and sample size while minimizing Euclidean distance to the utopian point (77 patients, 24 hours). This multi-objective methodology offers a transferable tool for managing competing priorities in wearable sensor data analysis, establishing a rigorous approach for multi-criteria decision-making that can be adapted across research domains. To our knowledge, no prior studies have applied multi-objective Pareto optimization specifically for temporal window selection with quality criteria in accelerometry/actigraphy data. While temporal window selection and Pareto optimization exist as a general concept, the systematic Pareto approach to balance competing objectives in psychiatric accelerometry is novel. In this study, Pareto optimization is employed as a search algorithm with two competing goals: maximizing patient inclusion and maximizing temporal window duration while maintaining data quality criteria. This multi-objective approach proves particularly advantageous in clinical research contexts compared to traditional single-objective search algorithms. Unlike conventional optimization methods that focus exclusively on identifying a single “best” solution, the Pareto framework systematically evaluates the entire solution space to identify non-dominated alternatives, providing clinical researchers with a comprehensive view of available trade-offs. This visualization of the complete decision landscape enables clinical personnel to make informed choices based on their specific research priorities, whether emphasizing larger sample sizes for statistical power or prioritizing data quality for signal integrity. Traditional search algorithms such as grid search, depth-first search (DFS), or breadth-first search (BFS) would converge to a single optimal point, obscuring alternative viable solutions that might better align with clinical constraints or research objectives. For larger search spaces involving more extensive datasets with increased patient populations and longer monitoring periods, incorporating advanced multi-objective algorithms such as NSGA-II (Non-dominated Sorting Genetic Algorithm II) could provide enhanced efficiency in exploring complex solution landscapes. However, in scenarios where only the single best approximate solution is required, classical approaches such as Genetic Algorithms or hyperheuristics might offer computational advantages while sacrificing the comprehensive decision-making transparency that makes Pareto optimization particularly valuable for clinical research applications.

After implementing FDR correction to account for multiple testing, none of the extracted motor activity features yielded statistically significant group differences between participants with ADHD and typically developing controls using Mann-Whitney U and Brunner-Munzel non-parametric tests within the optimal Pareto window. When using the same patients from the optimal Pareto window but with complete day data, none of the features were found to be significant. Only when using the uncleaned data did we find a significant feature: activity fragmentation with *p* < 0.01 in the Brunner-Munzel test after FDR correction. The Kruskal-Wallis statistical analysis for gender and age did not reveal any differences between the subgroups using the Pareto optimal window.

The three-dimensional Multidimensional Scaling (MDS) analysis provided compelling visual confirmation of our statistical findings. No clear clustering patterns emerged that distinguished the two diagnostic groups, as data points from both classes intermixed throughout the three-dimensional space in all temporal-quality configurations. Figure 3 corroborates the statistical inference results by demonstrating that when reducing the 34-feature space to its most informative three dimensions, the inherent structure of the data does not support class separation with the Pareto optimal window. The absence of distinct clusters or boundaries in the MDS plots (see repository) provides independent confirmation that the features lack sufficient discriminative power to distinguish ADHD from control patterns reliably.

**Figure 3.**
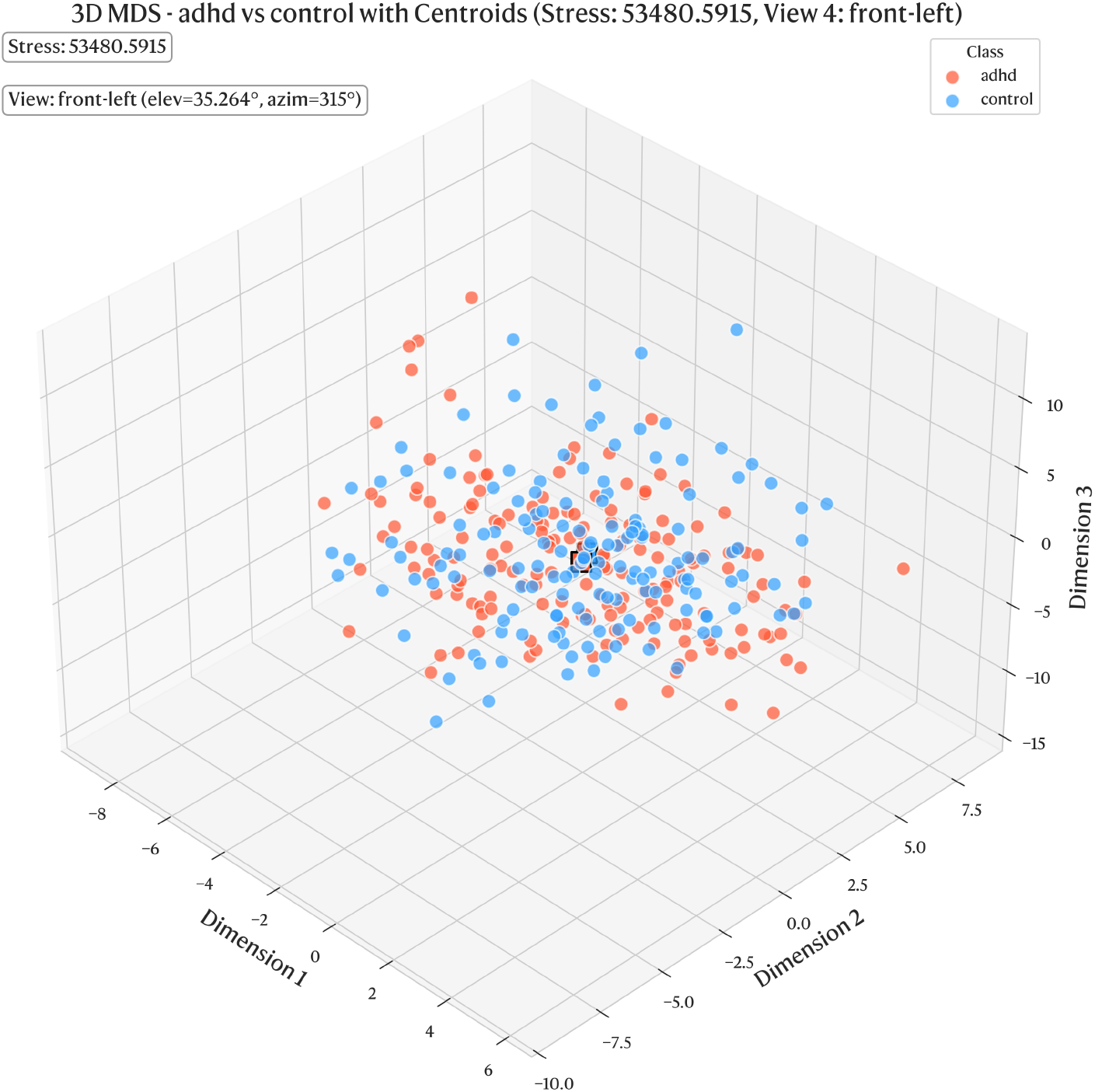
Multidimensional Scaling visualization showing the clustering capability of motor activity features in the optimal Pareto window configuration.

**Figure 4.**
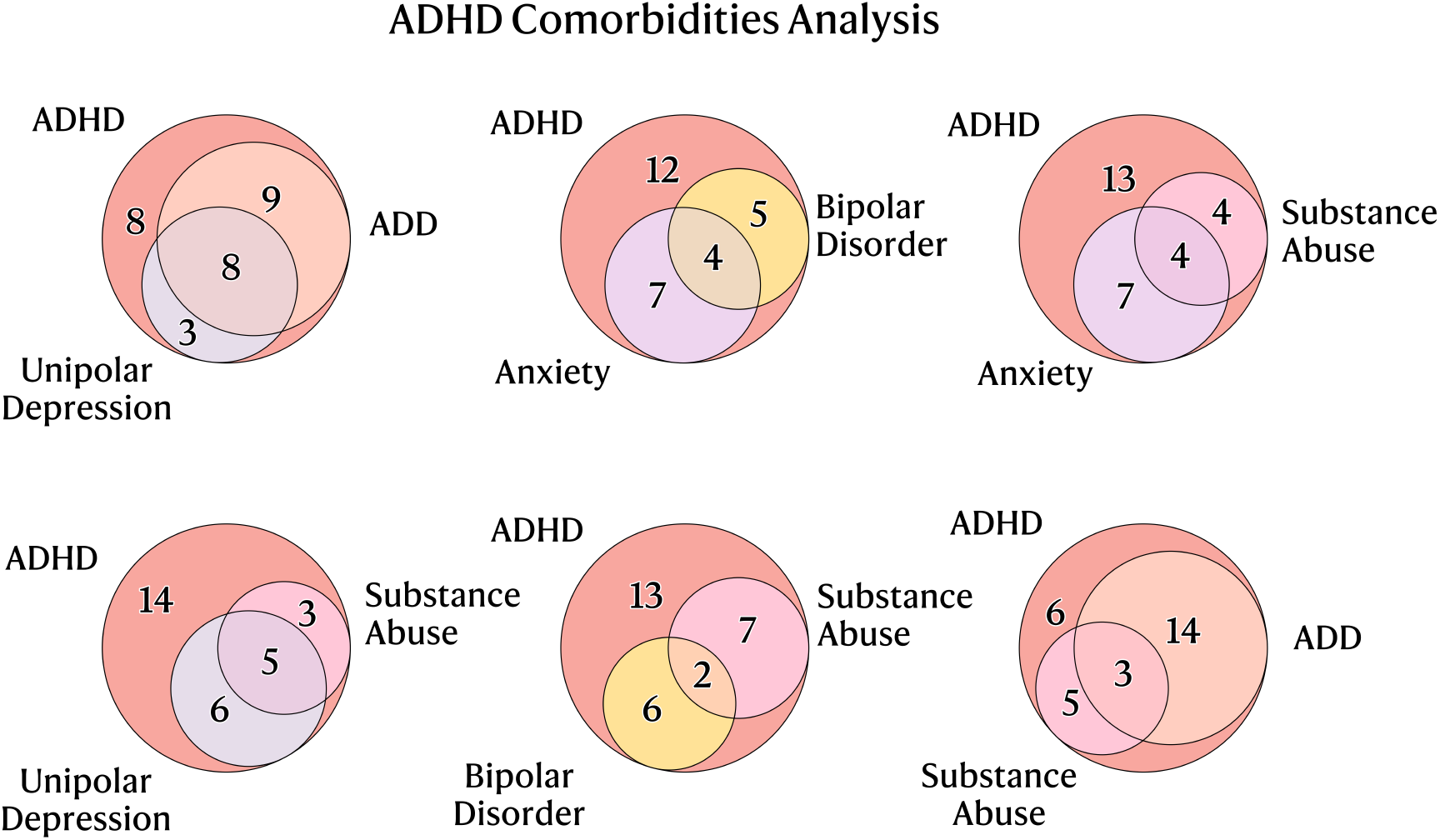
Distribution of psychiatric comorbidities in ADHD patients within the Pareto-optimal window (n=53).

**Figure 5.**
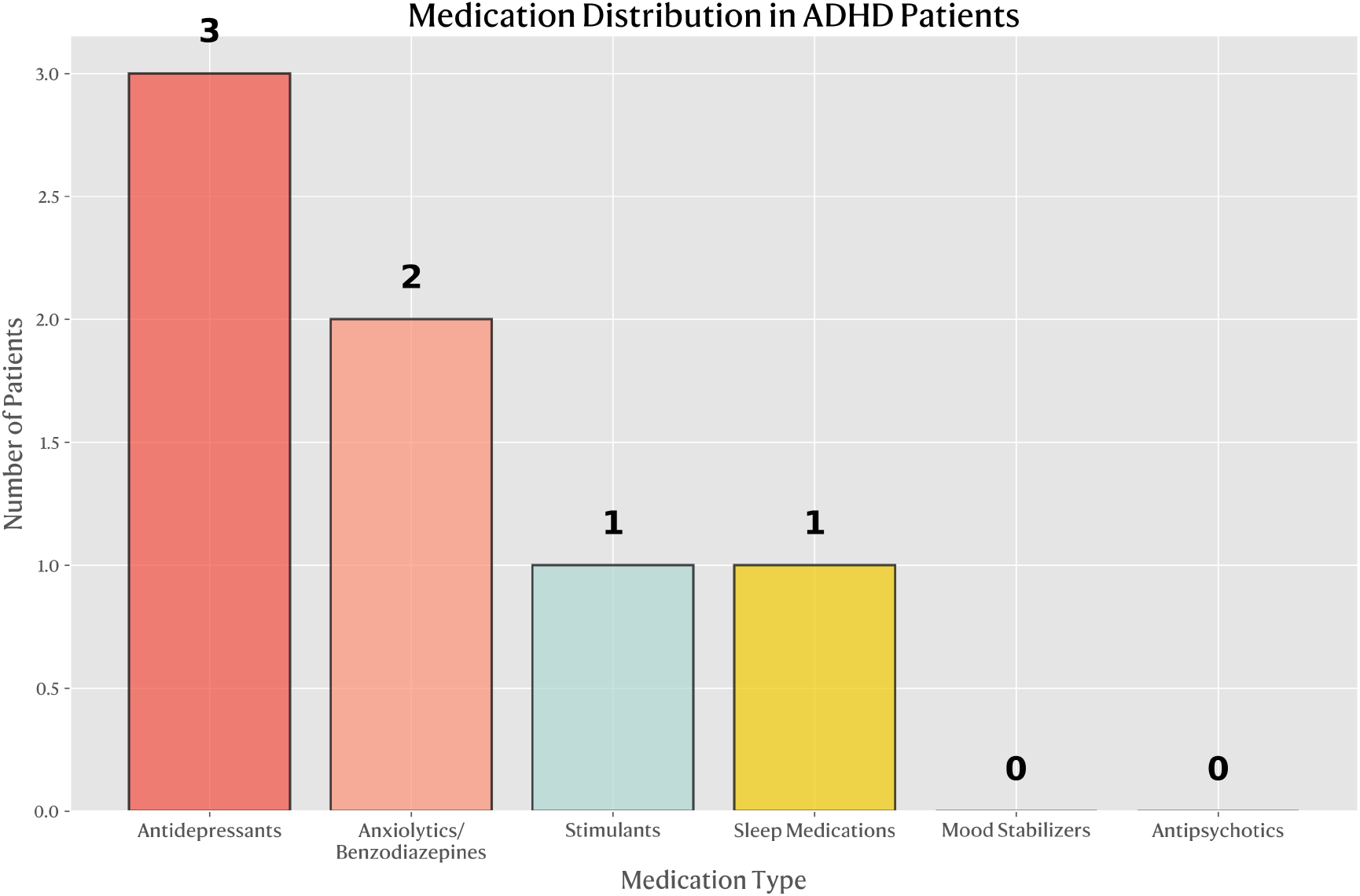
Medication usage patterns among ADHD patients in the Pareto-optimal window (n=53).

**Figure 6.**
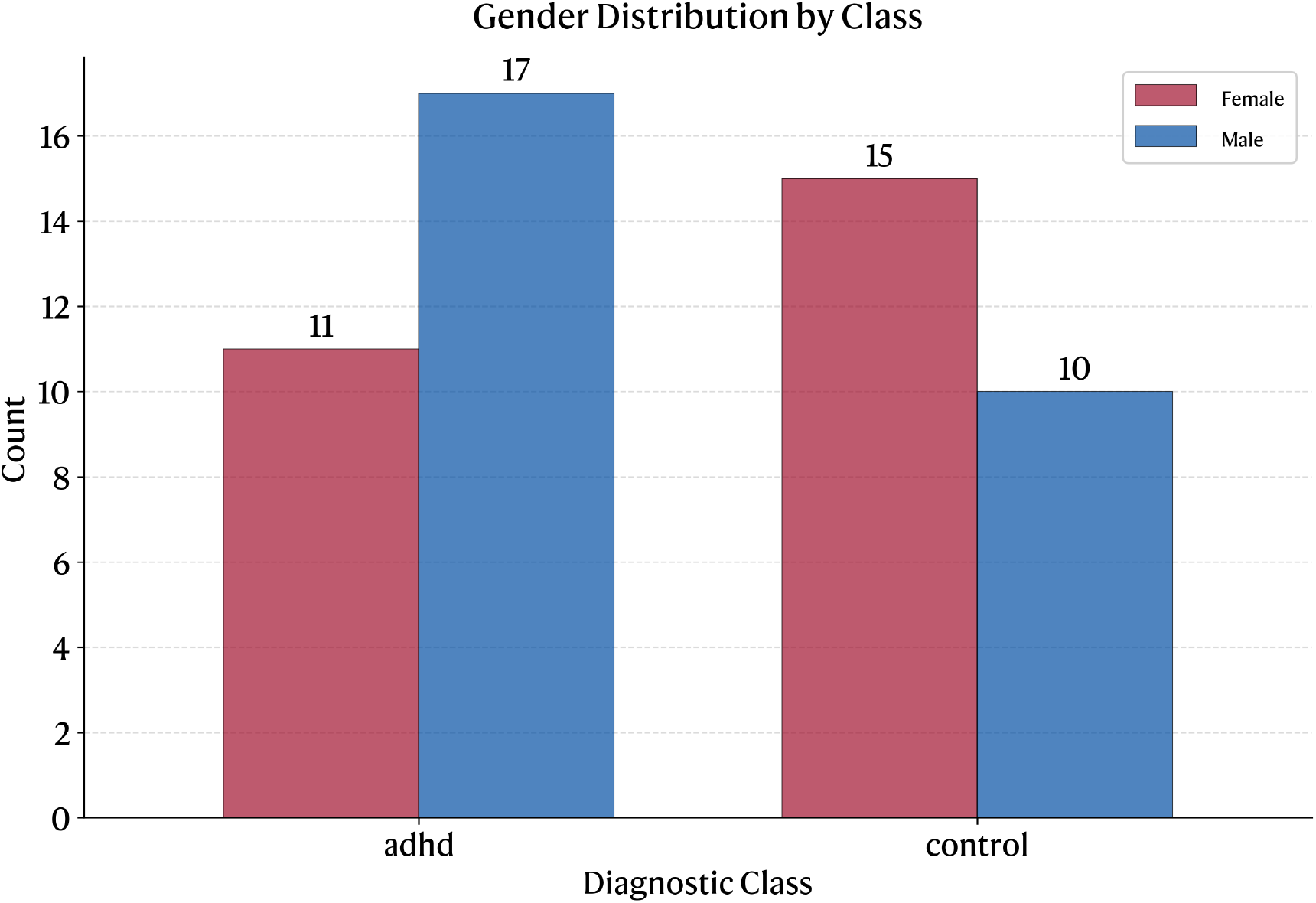
Gender distribution across ADHD and CONTROL groups in the Pareto-optimal window (n=53).

**Figure 7.**
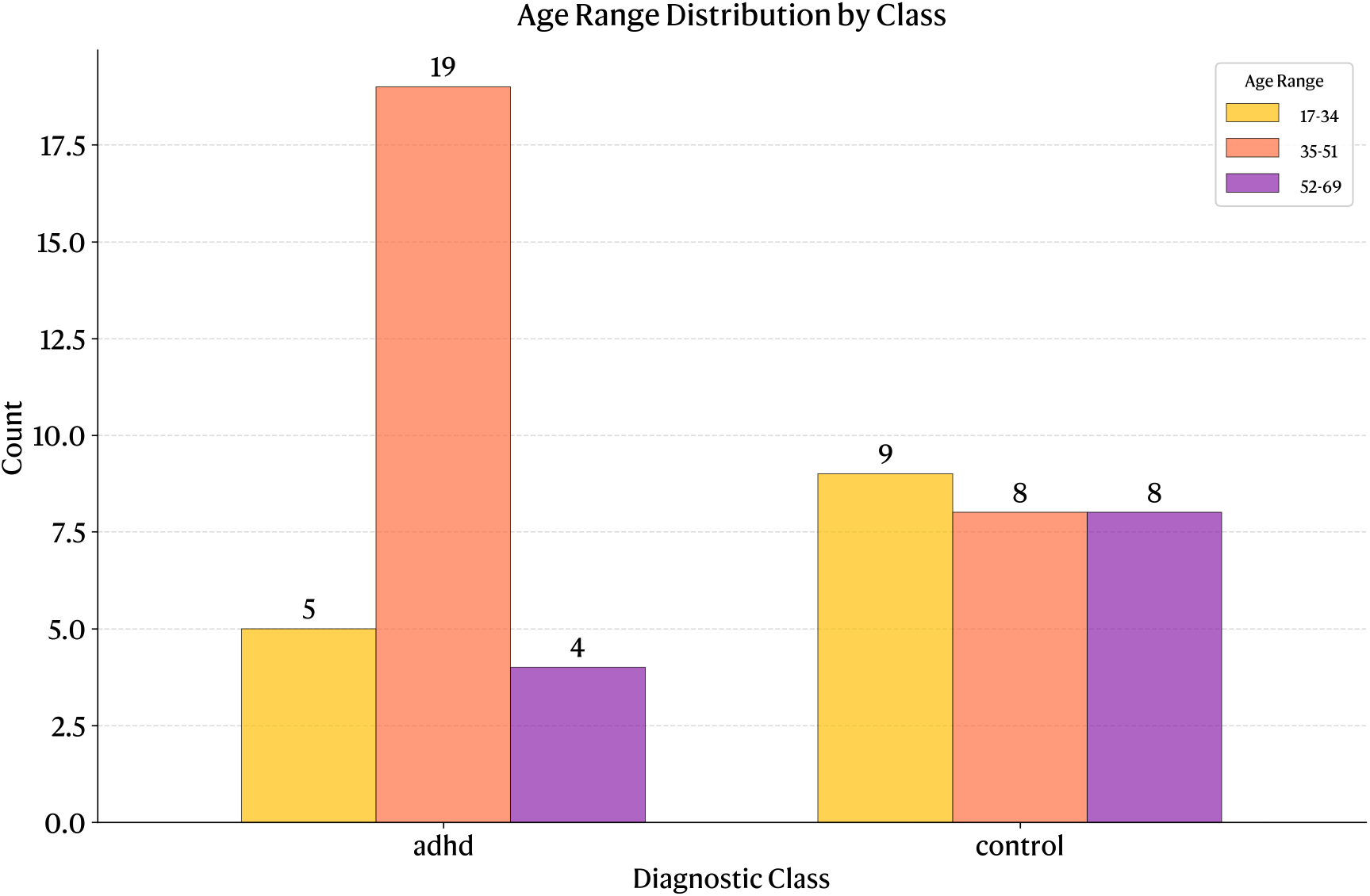
Age distribution across ADHD and CONTROL groups in the Pareto-optimal window (n=53).

The feature-to-feature correlation analysis, using Spearman’s rank correlation, revealed high multicollinearity among many extracted features, thereby reducing the effective dimensionality of the discriminative information. This redundancy explains, in part, why machine learning models struggled to identify unique discriminative patterns, as much of the apparent information content was duplicated across correlated features. Notably, the absence of significant differences persisted across all feature categories, from simple statistical measures to sophisticated complexity metrics and frequency domain analyses. This comprehensive result reinforces the conclusion that the lack of discriminative power is not due to inadequate feature engineering, but rather reflects the fundamental similarity between groups in their motor activity patterns when using a feature-based approach. This conclusion is consistent with Fasmer’s initial observations [45], which indicated no significant differences between the ADHD and CLINICAL groups over six days. However, it is crucial to recognize that the subjects and the criteria used to filter data in his study differ from those applied in this paper. Additionally, the observation that hyperactivity tends to decline with age [26] could provide a clinical rationale for the absence of significant differences when analyzing adults. However, the limitations of the dataset and the cross-sectional design inhibit definitive causal conclusions. Furthermore, among the ADHD patients in the original dataset, only seven presented with ADHD as an isolated condition without psychiatric comorbidities. This complicates the ability to confirm whether observed motor activity patterns are attributable to ADHD specifically rather than to other psychiatric conditions, such as unipolar depression, that may have predominant effects on motor behavior.

The machine learning evaluation across six traditional algorithms (Logistic Regression, Random Forest, Support Vector Machine, Multilayer Perceptron, K-Nearest Neighbors, and XGBoost) revealed a systematic pattern. Our analysis across three data quality configurations demonstrates how performance metrics degrade as data quality improves. With uncleaned data, as presented in Table II, Logistic Regression achieved a seemingly acceptable performance: 69% accuracy, 75% ROC AUC, and 37% Matthews Correlation Coefficient, with an ADHD precision of 76%, recall of 69%, and F1-score of 72% (Table III). With the same patients as the Pareto window, but considering their full six days, Logistic Regression performance degrades: accuracy drops to 62%, ROC AUC to 59%, and MCC to 24% (Table IV). With rigorous Pareto-optimal filtering, as shown in Table VI, all models converge to random performance, with the best performer in Table VII (MLP) achieving only 58% accuracy and 54% ROC AUC, essentially equivalent to random classification. This systematic degradation (ROC AUC: 75% → 59% → 54%) provides empirical proof that the uncleaned models were detecting systematic technical artifacts. More detailed results of these models, visualizations, and feature importance analysis can be found in the Zenodo repository [18]. The machine learning models employed a leave-one-out cross-validation strategy with majority voting at the daily level. This denotes that the patterns learned by the models were not generalizable across users, a finding that aligns with the high variability between subjects shown in the boxplots of the patient-level features.

**Table 2.** Results ML Complete Uncleaned ADHD CONTROL days Daily Features - Leave-One-Patient-Out Performance Summary.

| Model | Accuracy | ROC AUC | MCC |
| --- | --- | --- | --- |
| Dummy Most Frequent | 0.58 | 0.50 | 0.00 |
| Dummy Stratified | 0.56 | 0.50 | 0.09 |
| Dummy Uniform | 0.43 | 0.50 | -0.17 |
| Random Forest | 0.57 | 0.64 | 0.18 |
| SVM | 0.65 | 0.71 | 0.29 |
| MLP | 0.55 | 0.64 | 0.13 |
| Logistic Regression | **0.69** | **0.75** | **0.37** |
| XGBoost | 0.58 | 0.67 | 0.19 |
| KNN | 0.62 | 0.68 | 0.23 |

**Table 3.** Results ML Complete Uncleaned ADHD CONTROL days Daily Features - Logistic Regression Results (Best Model)

| Class | Precision | Recall | F1-Score | Support |
| --- | --- | --- | --- | --- |
| ADHD | **0.76** | 0.69 | **0.72** | 45 |
| CONTROL | 0.61 | 0.69 | 0.65 | 32 |
| Accuracy |  | **0.69** |  | 77 |
| Macro Average | **0.68** | 0.69 | **0.68** | 77 |
| Weighted Average | **0.70** | 0.69 | **0.69** | 77 |

**Table 4.** Results ML Pareto Complete Day Daily Features - Leave-One-Patient-Out Performance Summary.

| Model | Accuracy | ROC AUC | MCC |
| --- | --- | --- | --- |
| Dummy Most Frequent | 0.53 | 0.50 | 0.00 |
| Dummy Stratified | 0.57 | 0.50 | 0.12 |
| Dummy Uniform | 0.50 | 0.50 | 0.02 |
| Random Forest | 0.42 | 0.48 | -0.18 |
| SVM | 0.62 | 0.55 | 0.24 |
| MLP | 0.55 | 0.50 | 0.09 |
| Logistic Regression | **0.62** | **0.59** | **0.24** |
| XGBoost | 0.42 | 0.53 | -0.17 |
| KNN | 0.45 | 0.42 | -0.08 |

**Table 5.** Results ML Pareto Complete Day Daily Features - Logistic Regression Results (Best Model)

| Class | Precision | Recall | F1-Score | Support |
| --- | --- | --- | --- | --- |
| ADHD | **0.63** | 0.68 | **0.66** | 28 |
| CONTROL | 0.61 | 0.56 | 0.58 | 25 |
| Accuracy |  | **0.62** |  | 53 |
| Macro Average | **0.62** | 0.62 | **0.62** | 53 |
| Weighted Average | **0.62** | 0.62 | **0.62** | 53 |

**Table 6.** Results ML Pareto Trimmed Daily Features - Leave-One-Patient-Out Performance Summary.

| Model | Accuracy | ROC AUC | MCC |
| --- | --- | --- | --- |
| Dummy Most Frequent | 0.53 | 0.50 | 0.00 |
| Dummy Stratified | 0.57 | 0.50 | 0.12 |
| Dummy Uniform | 0.51 | 0.50 | 0.02 |
| Random Forest | 0.47 | 0.42 | -0.07 |
| SVM | 0.49 | 0.49 | -0.01 |
| MLP | **0.58** | 0.54 | **0.16** |
| Logistic Regression | 0.55 | 0.51 | 0.10 |
| XGBoost | 0.53 | **0.55** | 0.04 |
| KNN | 0.53 | 0.51 | 0.06 |

**Table 7.** Results ML Pareto Trimmed Daily Features - Multi-Layer Perceptron (MLP) Results (Best Model)

| Class | Precision | Recall | F1-Score | Support |
| --- | --- | --- | --- | --- |
| ADHD | **0.60** | 0.64 | **0.62** | 28 |
| CONTROL | 0.57 | 0.52 | 0.54 | 25 |
| Accuracy |  | **0.58** |  | 53 |
| Macro Average | **0.58** | 0.58 | **0.58** | 53 |
| Weighted Average | **0.58** | 0.58 | **0.58** | 53 |

A separate intra-patient variability analysis of the Pareto optimal window was conducted to assess the similarity between the features of different days from the same patient. The majority of the features remained stable, indicating that the days of the same patient were similar (coefficient of variation < 1.0). Only three features showed substantial variability: *intensity_gradient* (CV > 20 in ADHD versus ~ 2.5 in controls), *autocorrelation_90min* (CV ~ 8 in ADHD versus ~ 4 in controls), and *autocorrelation_30min* (CV ~ 2.5 in ADHD versus ~ 5). While ADHD showed slightly higher variability in specific features, these differences were modest and insufficient for reliable discrimination. Differences in mean temporal correlations were minimal (< 0.05 − 0.1) for most features, while individual patient variability (error bars) completely dominated any between-group differences. Only *dfa_scaling_exponent* and *sample_entropy* showed slightly different temporal correlations, but these remained within the range of individual variability. For the inter-patient consistency analysis via Intraclass Correlation Coefficients (ICC) of the majority of features, groups were remarkably similar (differences < 0.1), pointing to equivalent levels of internal homogeneity. Basic features like *mean* and *trimmed_mean* showed practically identical ICCs between groups. The singular notable exception was *activity_fragmentation*, which showed clearly lower ICC in ADHD (~ 0.37) versus controls (~ 0.75), suggesting greater inter-patient heterogeneity in fragmentation patterns among ADHD patients. This indicates that a monitoring approach with an individualized focus could be better suited to these data. High intra-patient daily feature similarity but high inter-patient variability could explain the poor classification performance of the models. Additionally, it may reinforce the conclusion that employing a K-fold validation at the daily level is inadequate, as it will mix days from the same patient between test and training splits, potentially leading the model to learn which patient is by similarity rather than because a discernible pattern exists across the classes. This separate variability analysis can be performed before deciding whether a machine learning approach is suitable, as it can provide a reasonable estimate of the expected behavior of the machine learning models.

The original protocol description for the ADHD study was obtained following communication with the Norwegian Regional Medical Research Ethics Committee West and consultation with the original principal investigator to facilitate a detailed analysis. The ADHD clinical protocol 251.08 underwent several modifications during its final implementation. A sleep diary was intended to be collected from each participant; however, due to incomplete data, it was ultimately excluded from the articles related to the dataset. The goal described in the protocol was to categorize ADHD participants into two distinct cohorts based on DSM-IV age criteria: one group comprised patients diagnosed according to standard age of onset, and the other included those diagnosed after the age of 7, alongside a third group of patients exhibiting minimal or no ADHD symptoms. The consensus diagnosis procedure involved four psychiatrists (KM, WF, OBF, and JØB) instead of the originally planned two (KM and WF). Although an inter-rater reliability assessment was attempted through videotaped interviews, it was deemed too time-consuming and consequently discontinued by the investigators. The protocol 150.01, which pertains to the PSYKOSE/DEPRESJON datasets from which the CONTROL group was derived, was not located by the committee or the original researchers and remains unavailable for analysis. From these changes, the principal limitation derived is the lack of information about each patient’s sleep patterns, which affects the interpretation of the accelerometry data. Additionally, the fact that the initial aim was to evaluate subgroups of the ADHD class suggests that variability in the activity patterns of the ADHD group was expected to be found depending on the age of onset, implying that a “one size fits all” approach may not accurately reflect motor activity in ADHD patients.

Given the substantial limitations in data quality, our findings should be interpreted as a quality assessment rather than a definitive statement about the clinical utility of accelerometry for detecting adult ADHD. Our findings that machine learning models demonstrate limited discriminative performance when rigorous data quality criteria are applied to the OBF-Psychiatric dataset represent a significant methodological contribution to the field. This result challenges assumptions about the direct applicability of machine learning approaches to clinical psychiatric classification with these data and underscores the critical importance of data quality assessment frameworks in clinical AI development. The development of standardized protocols for accelerometry data collection, processing, and quality assessment would facilitate meaningful comparisons across studies and reduce the risk of artifact exploitation. The results presented should not discourage accelerometry research for ADHD but rather highlight the need for appropriately powered validation studies using innovative methodological approaches.

## Limitations

Data collection was conducted using a device equipped with an omnidirectional accelerometer, which may not capture movement as accurately as tri-axis accelerometers can. The placement of the device on the wrist may have also constrained its ability to detect movements, such as continuous leg movement while sitting. The epoch used in this study is defined as a sum over a one-minute duration, which complicates direct comparisons with devices that utilize different settings, such as those that calculate epochs as means rather than sums. Additionally, there remains uncertainty regarding the extremely high acceleration values in the signals, as a count value between 7680 and 8000 per minute suggests an equivalent of 5g acceleration (128 counts) during all 60 seconds of the epoch. The metrological validation of the devices used to collect these accelerometry signals was not documented in the datasets descriptor articles or related previous works, suggesting this validation was not performed during the study.

There is no record of sleep hours, making it unlikely to determine with certainty which periods correspond to sleep time or to derive sleep metrics. Although patients received explicit instructions to wear the watch continuously and remove it only when bathing, we cannot verify which patients fully adhered to this protocol. Potential equipment damage, such as water exposure, might explain the unusual recordings observed during visual inspection of the actigraphy data. The signals contain extensive periods with zero count values. While this could indicate that the patient remained motionless or asleep during that time, it could also result from the watch being removed from their wrist. Furthermore, the device manual describes the sleep analysis functionality, where a value of at least 40 counts in a 1-minute epoch with medium sensitivity classifies a patient as awake [33]. This threshold increases the probability that extended periods with zero counts or values less than 40 may be due to not wearing the watch or sensor damage.

In terms of study design limitations, the application of strict quality exclusion criteria in this paper resulted in the absence of data for a specific day of the week for each person, as well as the loss of data from ADHD patients and control individuals. From a clinical and diagnostic perspective, it is essential to note that the standards for diagnosing ADHD have evolved since the DSM-IV criteria were utilized in this study, and our understanding of the disorder continues to expand. Moreover, there is currently no definitive framework for the assessment of adult ADHD patients based on motor activity patterns. Notably, the dataset descriptor does not report the assessment of potential medical conditions that could influence motor activity patterns or the presentation of ADHD symptoms, such as endocrinological conditions. Regarding sample characteristics, there were demographic imbalances, particularly in age and gender distribution between groups. While we found no significant differences between demographic subgroups in the features extracted, the limited sample size prevents definitive conclusions about model fairness and generalizability across demographic groups. The heterogeneity of ADHD, particularly in terms of comorbidities in the sample, adds further complexity. Substance abuse disorders and different medications in certain ADHD patients may also have impacted their behavioral patterns, complicating causal relationships. Our final small sample size (n=53) was determined by data availability rather than a priori power calculations, which represents a common challenge when working with pre-existing clinical datasets and cross-sectional designed studies. Also, this sample size complicates the ability to obtain a separate validation set distinct from the training and testing sets for hyperparameter tuning. No patients or members of the general public were involved in the design, conduct, or interpretation of this study, as we utilized a previously collected secondary dataset.

## Conclusion

Although the application of machine learning classifiers to accelerometry data has the potential to reveal patterns, the suboptimal data quality, small sample size, and confounding factors in these datasets limit the usefulness of classifiers in capturing the general characteristics of ADHD through accelerometry. Since model performance can vary significantly based on the training data used, preprocessing techniques, feature selection methods, validation strategies, and fixed hyperparameters, machine learning alone may introduce a layer of technical bias and subjectivity into the digital phenotyping of ADHD problems. Moreover, the substantial risk of overfitting in such small datasets requires a cautious interpretation of reported performance metrics. Importantly, despite having numerical observations, we cannot establish an objective view based on these data. Previous works published using the CONTROL or ADHD data from the OBF-Psychiatric dataset, or related ones (HYPERAKTIV, DEPRESJON, and PSYKOSE), require further evaluation.

This approach is deliberately reductionist, condensing the complex, non-stationary, and stochastic characteristics of continuous accelerometer signals into simplified daily summary statistics. Although this aggregation reduces temporal resolution and may hinder important within-day variability patterns, it provides practical benefits for initial classification and exploratory analysis, including lower computational requirements and enhanced interpretability. It is important to note that even with more sophisticated time series approaches, the same data confounding factors would persist. While these data can support many advanced numerical models, poor data quality will severely limit the effectiveness of any model.

Although we suggested a method for identifying a window that meets multiple criteria, signal quality should be prioritized from the outset of the data collection process. The quality filters applied in this paper are focused on this specific real-world case and are not ideal; they could be adapted or improved. Several technical constraints of the Actiwatch AW4, while present in our study, are less problematic in current accelerometry research. Modern devices, such as the GENEactiv [50], offer water resistance, circadian rhythm assessment, extended battery life, and enhanced data quality capabilities. The quality and viability of health machine learning models can also be addressed with TRIPOD-AI guidelines [51] and the recommendations for clinical predictive models discussed in Steyerberg’s book [52].

Fasmer’s visionary accelerometer research [45, 46] opens opportunities with modern devices and public discussion of accelerometry in psychiatric research. Understanding the data is the most valuable step we can take before deciding if a machine-learning approach is necessary. As researchers, we must maintain our focus; the primary objective of studying acceleration is to develop functional tools that assist clinicians and patients in identifying areas for improvement and training, rather than showcasing the best classifying machine learning algorithm with the highest accuracy or other performance metrics. Psychiatric and psychological research are more likely to have confounding factors than an “absolute typical norm”, but still, we have to try to approach the truth with the most detail possible to ensure traceability and aim to standardize procedures to provide reliable tools with reproducible tests.

## Data Availability

All data produced are available online at Zenodo.

https://doi.org/10.5281/zenodo.16396176

## Acknowledgment

This preprint was created using the LaPreprint template (https://github.com/roaldarbol/lapreprint) by Mikkel Roald-Arbøl https://orcid.org/0000-0002-9998-0058. AI assistance disclosure: Grammarly Pro was used for grammar and spelling corrections throughout the manuscript. Claude Sonnet 4 was used for language editing and sentence structure improvements. All scientific content, analysis, and conclusions remain the original work of the author.

## Notes

### Competing Interest Statement

The authors have declared no competing interest.

### Funding Statement

This study was funded by SECIHTI and Tecnologico de Monterrey indirectly by their support in P. Castillo-Gutierrez graduate scientific studies.

### Author Declarations

The study used only openly available human data that were originally located at: OBF-Psychiatric. DOI: 10.1038/s41597-025-04384-3.

